# Large-scale brain network analysis reveals functional-structural dissynchrony in HIV-associated asymptomatic neurocognitive disorders: Functional disturbances precede structural changes

**DOI:** 10.1101/2024.11.17.24317453

**Authors:** Zhongkai Zhou, Wenru Gong, Hong Hu, Fuchun Wang, Hui Li, Fan Xu, Hongjun Li, Wei Wang

**Author notes:** Corresponding authors: Wei Wang Hongjun Li. Zhongkai Zhou, Wenru Gong, and Hong Hu contributed equally to this work.

## Abstract

**Background:** In the era following combined antiretroviral therapy (cART), asymptomatic neurocognitive impairment (ANI) has become the primary stage of HIV-associated neurocognitive disorder (HAND). As a potentially reversible phase, precise identification of ANI is crucial. Multimodal MRI, with its non-invasiveness and high sensitivity, can reveal potential changes in brain network function and structure, providing significant support for exploring biomarkers of HAND and optimizing intervention strategies.This study aims to explore the dynamic changes in the functional network, structural network, and functional-structural coupling in ANI patients using multimodal MRI combined with large-scale brain network analysis.

**Methods:** A total of 95 participants were included, consisting of a healthy control group (HC, n=48) and an ANI patient group (n=47). Functional and structural connectivity matrices were constructed using resting-state fMRI (rs-fMRI) and diffusion tensor imaging (DTI), and graph theory analysis was used to evaluate global metrics, node characteristics, and functional-structural coupling changes.

**Results:** Structural Network: No significant changes were observed in the global or local topological properties of the structural network in ANI patients. Functional Network: Significant reorganization was observed in several key regions, including the visual network, executive control network, and default mode network. Functional-Structural Coupling: The functional-structural coupling in the occipital and frontal networks was significantly enhanced. Clinical Relevance: Changes in the functional network and functional-structural coupling were associated with the patients’ immune status, duration of infection, and cognitive performance.

**Conclusion:** The reorganization of the functional network and enhancement of functional-structural coupling during the ANI phase may reflect early manifestations of microscopic pathological changes (such as synaptic and dendritic damage). These changes hold promise as early warning signals in the progression of HAND and provide sensitive biomarkers and important research perspectives for precise diagnosis and early intervention.

## Introduction

Although combination antiretroviral therapy (cART) significantly suppresses HIV replication, bringing the life expectancy of people living with HIV (PLWH) close to that of the general population, a complete cure for HIV has not yet been achieved^[1, 2]^. HIV-associated neurocognitive disorders (HAND) are among the most common neurological complications, affecting approximately 18% to 47% of PLWH. These disorders are primarily characterized by deficits in attention, working memory, information processing speed, and fine motor skills^[3, 4]^. These deficits continue to impact the quality of life of individuals with HIV in areas such as medication adherence, financial management, driving ability, and employment, placing a heavy burden on families and society^[5, 6]^.

Based on the severity of cognitive impairment, HIV-associated neurocognitive disorder (HAND) is classified into asymptomatic neurocognitive impairment (ANI), mild neurocognitive disorder (MND), and HIV-associated dementia (HAD)^[7]^. With the effective viral suppression provided by cART regimens, the incidence of HAD has significantly declined. Currently, HAND primarily manifests as milder forms of cognitive impairment, particularly ANI^[8]^. Studies have shown that HAND is a potentially progressive condition, with ANI patients being 2 to 6 times more likely to develop symptomatic neurocognitive impairment compared to neuropsychologically normal (NP-N) individuals^[9, 10]^. As an early stage of HAND, ANI may be reversible^[11]^. Therefore, accurately identifying this stage and implementing early interventions is crucial for delaying or reversing the progression of HAND.

The current diagnostic criteria for HAND primarily rely on neuropsychological testing; however, these tests are often time-consuming and limited by subjectivity and operational environments, making it difficult to achieve early and accurate identification in clinical practice^[12]^. Therefore, identifying sensitive, reliable, and easily implementable imaging biomarkers to aid in the early diagnosis and intervention of HAND is of significant clinical importance. In recent years, multimodal MRI techniques and graph theory-based network analysis methods have provided important tools for studying the neuropathological mechanisms of HAND. The human brain is viewed as a complex large-scale network, where specific connection patterns enable local processing and global integration^[13]^. Structural connectivity networks reflect anatomical connections between brain regions, while functional connectivity networks capture dynamic interactions. Together, they exhibit the small-world properties of the brain that optimize information processing^[14, 15]^. Furthermore, structural connectivity provides the physical foundation for functional connectivity, which, through plasticity mechanisms, reciprocally influences structural connectivity. The dynamic coupling of these two networks is crucial for maintaining brain function^[16–18]^. Existing multimodal MRI studies have shown persistent deficits in the structure, function, and network connectivity of the brains of HIV-infected individuals^[19–23]^. However, the specific patterns of brain function and structural network changes during the early ANI phase of HAND have not been fully elucidated. There remains a lack of exploration, particularly regarding the functional-structural coupling relationships at different network levels. Moreover, significant discrepancies in existing research findings limit a comprehensive understanding of the early neuropathological mechanisms of HAND^[24–27]^.

To further elucidate the neuropathological features of early HAND and provide more sensitive and objective imaging evidence for its early diagnosis and intervention, this study uses multimodal MRI data combined with large-scale brain network analysis to systematically investigate the functional and structural network changes in ANI patients. Based on rs-fMRI and DTI data from the same cohort, functional connectivity networks (FCN) and structural connectivity networks (SCN) are constructed. Through graph theory analysis, this study is conducted in the following four aspects: (1) Systematically analyze the topological structural features of FCN and SCN in ANI patients to investigate whether there is common or differential damage; (2) Explore the changes in functional-structural coupling at the whole-brain and subnetworks levels in ANI patients to reveal potential coupling dysregulation patterns; (3) Use network-based statistical analysis methods to detect abnormal regions in functional or structural connectivity in ANI patients; (4) Evaluate the correlation between changes in network topology and clinical variables and cognitive performance in patients.

## Materials and Methods

### Participants

This study was approved by the Medical Ethics Committee of Beijing You’an Hospital, Capital Medical University. All participants provided written informed consent in accordance with the Declaration of Helsinki (Ethics approval number: LL-2020-047-K). According to the Gisslén criteria^[28]^, from November 2020 to April 2024, this study recruited 47 individuals with HIV in the asymptomatic neurocognitive impairment (ANI) phase and 48 healthy controls (HC) who were confirmed to be free of HIV-associated neurocognitive disorders (HAND) through neuropsychological (NP) testing at the outpatient clinic of the Infectious Disease Center at Beijing You’an Hospital. The healthy control group was matched to the patient group based on age, gender, and education level. Inclusion criteria for HIV patients were as follows: (1) Asian ethnicity (Han Chinese); (2) age between 20 and 60 years; (3) right-handed. Exclusion criteria included: (1) presence of central nervous system (CNS) tumors, infections, cerebrovascular diseases, or other systemic illnesses; (2) history of neurological or psychiatric disorders, such as anxiety or depression; (3) history of alcohol or drug abuse; (4) contraindications for MRI scans. Demographic and clinical laboratory data were obtained from electronic health records (EHR), including age, gender, education level, duration of HIV diagnosis, duration of cART treatment, lowest CD4^+^ cell count after HIV infection, plasma CD4^+^ cell count within two weeks prior to NP assessment, CD4^+^/CD8^+^ ratio, and current plasma viral load.

### Neuropsychological test

According to the Gisslén criteria, the assessment should cover at least five cognitive domains, and ANI is defined as performance more than 1.5 standard deviations below the age-and education-adjusted normative score mean in at least two cognitive domains^[28]^. This study assessed five cognitive domains in people living with HIV (PLWH) through a set of neurocognitive tests standardized for age, gender, education level, and residential area size^[29]^. These cognitive domains included: (1) speed of information processing [Trail-Making Test Part A (TMT A)]; (2) memory, including learning and recall [Hopkins Verbal Learning Test-Revised (HVLT-R) and Brief Visuospatial Memory Test-Revised (BVMT-R)]; (3) attention and working memory [Continuous Performance Test-Identical Pairs (CPT-IP), Wechsler Memory Scale-III (WMS-III), and Paced Auditory Serial Addition Test (PASAT)]; (4) fine motor skills [Grooved Pegboard Test]; and (5) verbal and language skills [Animal Naming Test].

### Data acquisition

Structural, functional, and diffusion tensor imaging (DTI) data for all participants were acquired using a Siemens 3.0 T magnetic resonance imaging (MRI) scanner (Siemens Trio Tim B17 software, Germany) with a 32-channel dedicated head coil. Prior to scanning, headrests and neck supports were adjusted to ensure the participants were comfortably lying supine on the examination table, with the head secured using a latex pad to minimize head movement during the acquisition process. Before image acquisition, researchers communicated with participants to ensure they were in a relaxed state. Participants were allowed to close their eyes but were required to remain awake and wore earplugs or headphones to reduce noise interference from the scanning equipment. T1-weighted structural imaging was performed using a magnetization-prepared rapid gradient echo sequence (MP-RAGE) with the following scanning parameters: repetition time (TR) = 1900 ms, echo time (TE) = 2.52 ms, inversion time (TI) = 900 ms, acquisition matrix = 256 x 246, field of view (FOV) = 250 x 250 mm², flip angle = 9°, voxel size = 1 x 0.9766 x 0.9766 mm³, and acquisition time of 4 minutes and 18 seconds. Resting-state fMRI (Rs-fMRI) was performed using a gradient echo single-shot echo planar imaging sequence (GE-EPI) with the following scanning parameters: TR = 2000 ms, TE = 30 ms, acquisition matrix = 64 x 64, voxel size = 3.5 x 3.5 x 4.2 mm³, flip angle = 90°, 35 slices, 240 time points, with an acquisition time of 8 minutes and 6 seconds. Diffusion tensor imaging (DTI) was performed using a spin-echo echo planar imaging sequence (SE-EPI) with the following scanning parameters: TR = 9200 ms, TE = 85 ms, slice thickness = 2 mm, no gap, 65 slices, matrix size = 112 x 112, FOV = 224 x 224 mm², number of excitations = 1, spatial resolution = 2.0 x 2.0 x 2.0 mm³, total acquisition time = 10 minutes and 27 seconds, and phase encoding direction = anterior-posterior (AP). Diffusion-sensitized gradients were applied along 64 non-collinear directions, with b-values of 1000 s/mm² and 0 s/mm².

### Brain network construction

#### Anatomical parcellation

Functional connectivity MRI studies have shown that the cerebellum projects to multiple brain networks and limbic regions through the cortico-ponto-cerebellar and cerebello-thalamo-cortical circuits, leading to impairments in executive function, spatial cognition, language abilities, and changes in personality and emotions^[30, 31]^. Structural MRI has also confirmed that HIV infection can lead to significant atrophy of the cerebellar cortex^[32]^. Therefore, in this study, to determine the nodes of the whole-brain functional and structural connectivity networks, we used the AAL116 brain atlas^[33]^. The AAL116 atlas divides the entire brain (including the cerebellum) into 116 anatomical regions of interest (ROIs), with 90 regions located in the cortical and subcortical structures, and 26 regions in the cerebellum. A list of the anatomical labels for the nodes can be found in the supplementary materials, as shown in Table S1.

### Functional connectivity network construction

The Rs-fMRI data were preprocessed and matrix construction was performed using RESTplus 1.25 software^[34]^, based on Statistical Parametric Mapping (SPM 12, https://www.fil.ion.ucl.ac.uk/spm/), and executed on the Matlab 2022b platform (MathWorks, Natick, MA, USA). The main steps included: (1) converting the raw MRI images to the Neuroimaging Informatics Technology Initiative (NIFTI) format for subsequent processing; (2) Removing the first 10 time points from the 240 sampling points to correct for temporal shifts within volumes and geometric displacements between volumes; (3) Temporal slice timing correction; (4) Head motion correction, ensuring no participant’s displacement in any direction exceeded 3 mm or angular rotation exceeded 3°; (5) Spatial normalization was performed using the DARTEL algorithm with an extended exponential Lie algebra deformation approach^[35]^, followed by resampling to 3 x 3 x 3 mm³; (6) Removal of linear trends from the time series; (7) Regression of confounding covariates, including Friston-24 head motion parameters, global brain signal, white matter signal, and cerebrospinal fluid signal^[36]^; (8) Bandpass filtering within the range of 0.01–0.08 Hz. To obtain the functional connectivity of each participant, the 116 ROIs defined by the AAL116 atlas were considered as nodes, and the Pearson correlation coefficient between the time series of each pair of nodes was calculated to generate the functional connectivity correlation matrix (r matrix). Given the uncertainty regarding the biological interpretation of negative correlations^[37]^, the analysis was restricted to positive correlations, with negative correlation coefficients set to zero. Finally, an undirected weighted network was generated for each participant. Additionally, the r matrix was transformed to a z matrix using Fisher’s Z transformation to improve normality.

### Structural connectivity network construction

The DTI data were first converted from DICOM format to NIFTI format using MRIcroGL software, and then the whole-brain fiber number (FN) connectivity matrix was constructed using DSI Studio software (https://dsi-studio.labsolver.org/). The specific steps are as follows: (1) Load the NIFTI format images and create SRC files; (2) Use an automatic quality control program to check the b-table, ensuring the accuracy of diffusion directions and b-values; (3) Confirm the white matter region mask coverage to improve the reconstruction quality. Next, head motion and eddy current distortion correction were performed, and image reconstruction was carried out using the generalized q-sampling imaging (GQI) method. The reconstructed images were then registered to the ICBM152 standard space, resulting in the generation of FIB files; (4) Load the FIB files and perform fiber tracking using a deterministic fiber tracking algorithm, removing fiber paths shorter than 20 mm or longer than 200 mm, with a threshold of 1 million fibers as the endpoint limit. The brain parcellation was performed using automatic anatomical labeling software, dividing the brain into 116 nodes, consistent with the rs-fMRI images. The number of fiber bundles between these nodes was calculated to construct an undirected unweighted FN matrix, representing the whole-brain structural connectivity network.

### Network analysis

Graph theory analysis was performed on the HC group and ANI patient group using the Brain Connectivity Toolbox GRETNA (https://www.nitrc.org/projects/gretna/)^[38]^. First, the current r matrix and FN matrix were each transformed into binary undirected 116 x 116 network matrices using sparsity thresholding and element value thresholding. Subsequently, the global and nodal network properties of the functional connectivity network (FCN) and structural connectivity network (SCN) were calculated.

### Global topologic parameters

The global attributes of this study primarily focus on key metrics of FCN and SCN, including small-world properties, global efficiency, and modularity. The small-world properties include the clustering coefficient (Cp), characteristic path length (Lp), normalized clustering coefficient (γ), normalized characteristic path length (λ), and small-worldness index (σ)^[39]^. Specifically, γ represents the ratio of the network’s clustering coefficient to that of a random network, λ represents the ratio of the network’s characteristic path length to that of a random network, and σ is calculated as the ratio of γ to λ (σ = γ/λ). To ensure that the FCN of all participants meets the small-world property criterion (σ > 1.1), we used the Markov-chain algorithm to generate 1000 random networks^[40]^ To determine the lower limit of the sparsity threshold, the function [a, b, c] = gretna_get_rmax(rand(116)) was used for computation, with a step size set to 0.01. The upper limit of sparsity was selected under the condition of σ > 1.1, thereby combining the absolute connectivity strength threshold with the relative sparsity threshold^[41]^. The global efficiency metrics include global efficiency (Eg) and local efficiency (Eloc)^[42]^, where Eg assesses the overall effectiveness of the network in parallel information transmission, while Eloc quantifies the network’s local information transmission capacity, partly reflecting the network’s robustness against external disruptions. Modularity is used to divide the whole-brain network into multiple subnetworks, where nodes within each module are more densely connected, while connections between modules are relatively sparse^[43]^. Given that this study analyzes both FCN and SCN, the whole-brain network was divided into seven subnetworks based on anatomical regions: frontal lobe, prefrontal lobe, subcortical, parietal lobe, temporal lobe, occipital lobe, and cerebellum network^[44, 45]^.

### Regional nodal characteristics

This study calculated six node metrics, including degree centrality (Dc), betweenness centrality (Bc), nodal clustering coefficient (NCp), nodal efficiency (Ne), nodal local efficiency (NLe), and nodal shortest path length (NLp). Specifically, Dc represents the number of connections a node has with other nodes, reflecting the direct influence of that node in the network; Bc measures the frequency with which a node serves as an intermediary in the shortest paths between other nodes, indicating its key mediating role in the network; NCp evaluates the degree of interconnection between a node’s neighbors, reflecting the local clustering of the network. Its value is the ratio of the actual number of connections to the maximum possible number of connections; Ne represents the information transmission efficiency from a node to all other nodes, typically determined by calculating the inverse of the shortest path length between the node and other nodes; NLe measures the communication efficiency that can still be maintained between neighboring nodes when a node fails, thus reflecting the network’s local fault tolerance; NLp represents the average shortest path length from a node to all other nodes, reflecting the average distance of that node within the network^[39, 46–48]^.

### Network connectivity characteristics

This study employed z-matrices and FN matrices, using the Network-Based Statistic (NBS) method to conduct a localization analysis of the connectivity changes between brain regions in FCN and SCN. The NBS method controls the familywise error rate through large-scale univariate testing. Its underlying principle is similar to traditional cluster-based statistical parametric mapping thresholding methods^[49]^.

### Coupling between functional and structural connectivities

The coupling between functional connectivity (FC) and structural connectivity (SC) in each participant was investigated, with a focus on the whole-brain connectivity level and modular level. (1) Whole-brain connectivity level: The coupling between FC (the Fisher’s Z-transformed version of the Pearson correlation coefficient) and SC was quantified by calculating the correlation between the FC and SC matrices. The correlation between functional and structural connectivity is constrained by the presence of non-zero structural connections^[50]^. First, non-zero structural connections were extracted to generate a structural connectivity vector, and these values were resampled to follow a Gaussian distribution with a mean of 0.5 and a standard deviation of 0.1^[16]^. Next, the corresponding functional connectivity vector was extracted, and the Pearson correlation coefficient between the two vectors was calculated. (2) Focused on the differential modules identified in the functional and structural modularity analyses. For each differential module, non-zero structural connections within and between the modules were extracted to generate the structural connectivity vector, and the corresponding functional connectivity vector was also extracted. Using the same method as for the whole-brain connectivity level, the Pearson correlation coefficient between these two vectors was computed to obtain the coupling value of the differential modules.

### Statistical analysis

Statistical analysis of clinical demographic data and cognitive performance between the HC group and ANI patient group was conducted using IBM SPSS Statistics 29 (IBM Corp., Armonk, NY, USA). Categorical variables were expressed as frequencies (percentages), and chi-square tests were used to compare inter-group differences. The normality of continuous variables was assessed using the Shapiro-Wilk test. Variables with normal distribution were expressed as means ± standard deviations (*x̅* ± s), and variables not following a normal distribution were presented as medians (interquartile ranges) [*M(IQR)*]. Inter-group differences were compared using t-tests or Mann-Whitney U tests. The significance level was set at *P* < 0.05.

To assess global properties such as small-world characteristics, whole-brain efficiency, and modularity, as well as node-level attributes, two-sample *t*-tests were conducted using GRETNA to compare inter-group differences in network metrics. In modularity and node-level attribute analyses, Bonferroni correction for multiple comparisons (*P* < 0.05) was applied to control the false-positive rate.

For inter-group differences in functional and structural network connectivity, Network-Based Statistic (NBS) analysis was performed using GRETNA. First, two-sample t-tests were conducted to compare inter-group network connectivity differences, with an initial threshold set at *P* < 0.001 to identify supra-threshold connections^[51]^. Subsequently, the P-value threshold in the permutation tests was set at 0.05 to control the significance level of sub-networks forming clusters. A total of 5000 permutation tests were conducted to generate an empirical null distribution based on random group membership to evaluate the statistical significance of each observed component size.

Two-sample t-tests and visualizations of functional-structural connectivity coupling were performed using GraphPad Prism 10.1.2 (https://www.graphpad-prism.cn/), with the significance level set at *P* < 0.05.

Furthermore, to explore the relationship between network topology changes in ANI patients and clinical variables and cognitive performance, correlation heatmaps were generated using the corrplot package in RStudio (https://posit.co/products/open-source/rstudio/). The heatmap displayed the Pearson correlation coefficients between variables, with color intensity representing the strength of the correlations. During the correlation analysis, False Discovery Rate (FDR) correction was applied to control for multiple comparisons. The significance level was set at *P* < 0.05.

## Results

### Demographic data and cognitive performance

No significant differences were observed between the HC group and ANI patient group in terms of age, gender, and educational level (all P > 0.05). As expected, ANI patients performed significantly worse than the HC group across all cognitive domains (i.e., speed of information processing, memory (learning and recall), attention/working memory, fine motor skills, verbal and language) (all P > 0.05). The demographic data and cognitive performance of the 95 participants are shown in Table S2.

### Disrupted functional connectivity network

Both the HC and ANI groups exhibited small-world characteristics in the functional network, with σ > 1.1, and the sparsity upper limit was set to 0.5. Statistical analysis revealed that, compared to HC, ANI patients exhibited significantly increased σ (*P* = 0.032) and Eg (*P* = 0.005), while Cp (*P* = 0.004), Lp (*P* = 0.007), λ (*P* = 0.009), and Eloc (*P* = 0.005) were significantly decreased. However, no significant difference was observed in γ between the two groups (*P* = 0.735), as shown in Figure 1 and Table S3. Modular analysis showed that, within the anatomically defined 7 subnetworks, the functional connectivity within the occipital network module was significantly enhanced in ANI patients (*P* < 0.001, T = -3.472, Bonferroni correction). Additionally, ANI patients exhibited significantly reduced intermodular functional connectivity between the prefrontal network and the cerebellar network (*P* = 0.002, T = 3.145, Bonferroni correction), as shown in Figure 2.

**Fig 1.**
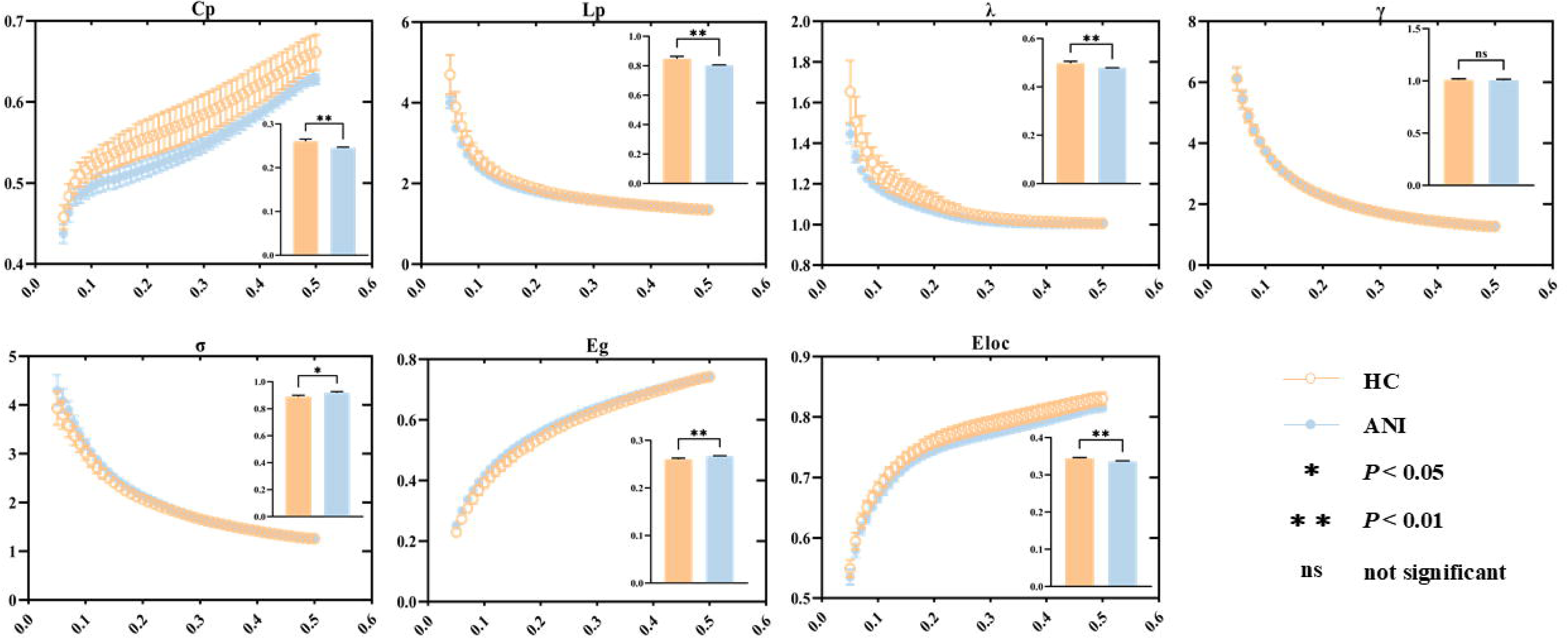
The functional connectivity network’s global properties and their functional relationship with various sparsity thresholds. The embedded bar chart displays the mean area under the curve for each metric, with vertical bars indicating the standard error between subjects. * *P* < 0.05, ** *P* < 0.01. Abbreviations: HC, healthy control; ANI, asymptomatic neurocognitive impairment; Cp, clustering coefficient; Lp, characteristic path length; λ, normalized characteristic path length; γ, normalized clustering coefficient; σ, small-world topology; Eg, global efficiency; Eloc, local efficiency; ns, not significant.

**Fig 2.**
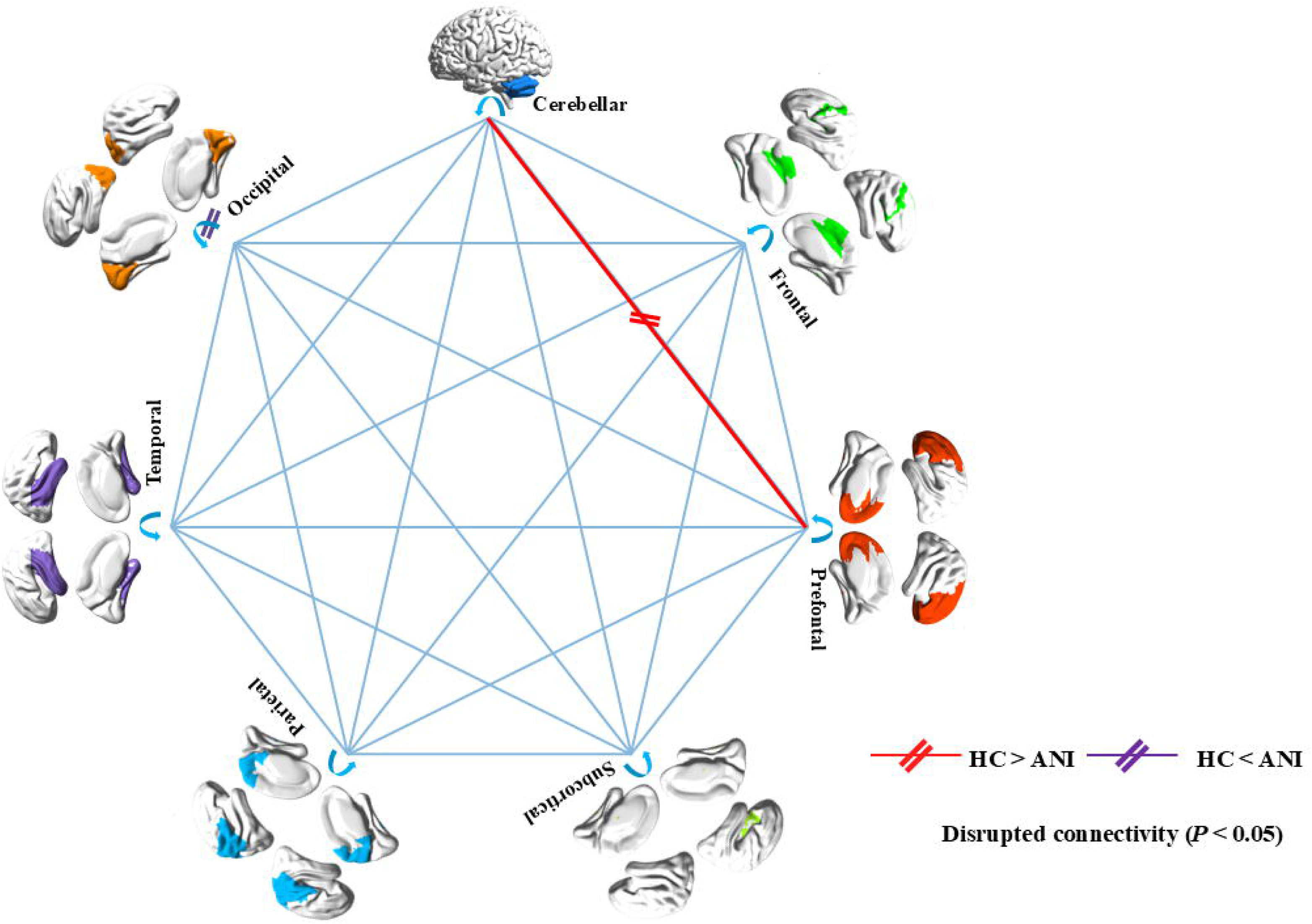
Changes in intra-and inter-module functional connectivity associated with ANI (*P* < 0.05, Bonferroni corrected). The wide lines represent significant T-values from between-group comparisons (red indicates HC > ANI, purple indicates HC < ANI).

In node-level analysis, compared to HC, ANI patients showed a significant reduction in Dc in the left inferior orbital gyrus (ORBinf.L) (*P* < 0.001, Bonferroni correction), whereas Dc was significantly increased in the right calcarine sulcus surrounding cortex (CAL.R), left superior occipital gyrus (SOG.L), left precuneus (PCL.L), and right precuneus (PCL.R) (all *P* < 0.001, Bonferroni correction). The nodal clustering coefficient (NCp) was significantly reduced in the left olfactory cortex (OLF.L), left putamen (PAL.L), and right putamen (PAL.R) (all *P* < 0.001, Bonferroni correction). Ne was significantly reduced in the left calcarine sulcus surrounding cortex (CAL.L), right calcarine sulcus surrounding cortex (CAL.R), left lingual gyrus (LING.L), right lingual gyrus (LING.R), left superior occipital gyrus (SOG.L), left middle occipital gyrus (MOG.L), left inferior occipital gyrus (IOG.L), left precuneus (PCUN.L), right precuneus (PCUN.R), left central precuneus (PCL.L), right central precuneus (PCL.R), and left cerebellar sixth lobe (CRBL6.L) (all *P* < 0.001, Bonferroni correction). The nodal local efficiency (NLe) in the left caudate nucleus (CAU.L) was significantly reduced (*P* < 0.001, Bonferroni correction). The nodal shortest path length (NLp) was significantly reduced in the left lingual gyrus, right lingual gyrus, and left cerebellar sixth lobe (CRBL6.L) (all *P* < 0.001, Bonferroni correction). Additionally, no significant differences in betweenness centrality (Bc) were found between the two groups, as shown in Figure 3 and Table S4.

**Fig 3.**
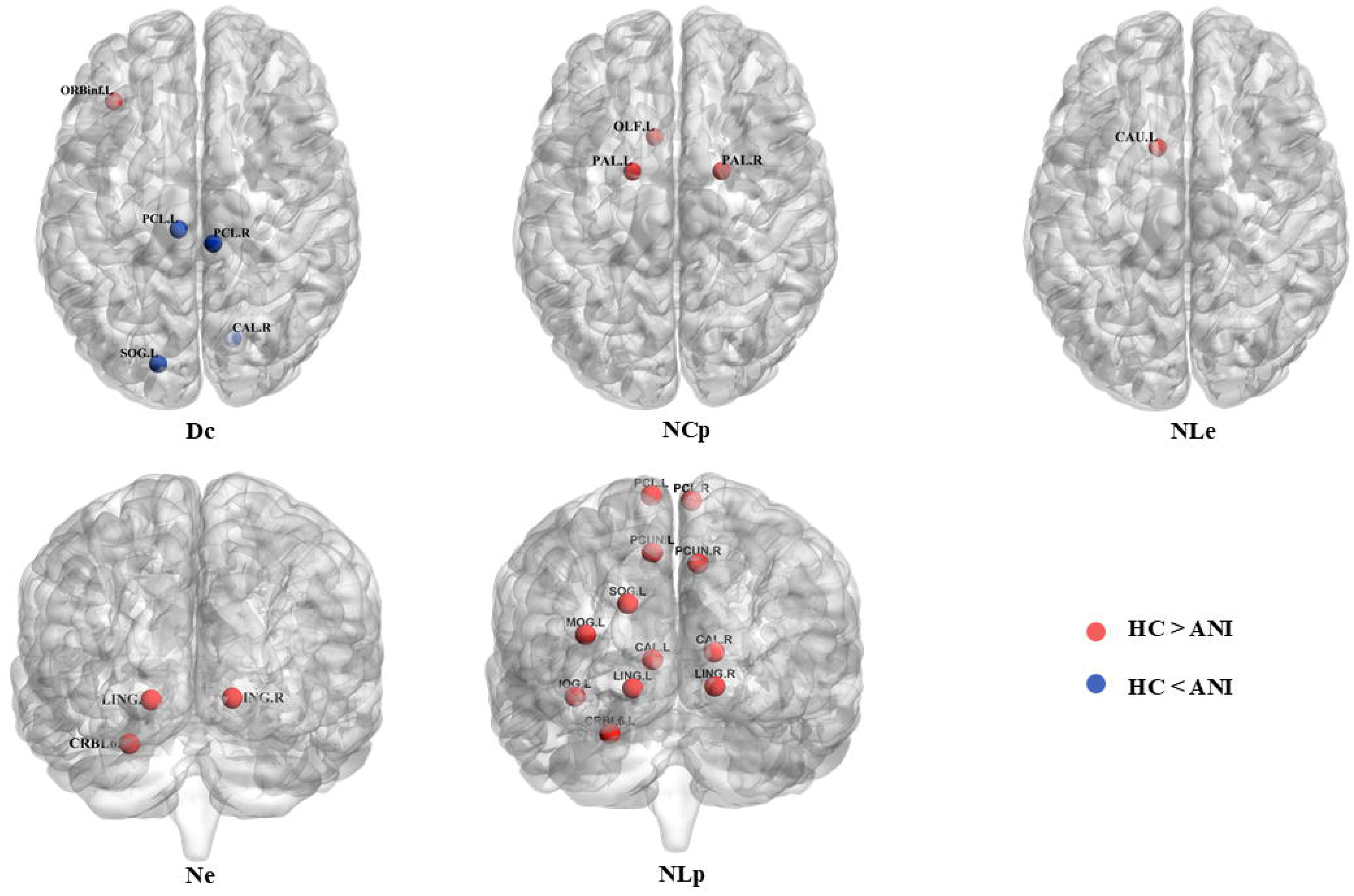
Significant changes in the node properties of the functional connectivity network in ANI patients relative to HC, using the AAL-116 atlas (*P* < 0.05, Bonferroni correction). Red indicates HC > ANI, and Klein blue indicates HC < ANI. Abbreviations: Dc, degree centrality; NCp, nodal clustering coefficient; NLe, nodal efficiency; Ne, nodal efficiency; NLp, nodal shortest path length; ORBinf.L, left orbitofrontal inferior gyrus; CAL.R, right periaqueductal fissure cortex; SOG.L, left supraoccipital gyrus; PCL.L, left paracentral lobule; PCL.R, right paracentral lobule; OLF.L, left olfactory cortex; PAL.L, left pallidum; PAL.R, right pallidum; CAL.L, left periaqueductal cortex; CAL.R, right periaqueductal cortex; LING.L, left lingual gyrus; LING.R, right lingual gyrus; SOG.L, left supraoccipital gyrus; MOG.L, left middle occipital gyrus; IOG.L, left inferior occipital gyrus; PCUN.L, left precuneus; PCUN.R, right precuneus; CRBL6.L, left cerebellar lobe VI; CAU.L, left caudate nucleus.

### Stable Structural Connectivity Metrics

Surprisingly, in terms of structural topology, no significant differences were observed between the two groups in the global metrics, namely Cp, Lp, γ, λ, σ, Eg, and Eloc (all *P* > 0.05) (Figure S1, Table S5). Furthermore, in the modularity analysis, no significant differences were found in the modular structural connectivity both between and within the 7 subnetworks (all *P* > 0.05, Bonferroni correction). In the node-level analysis, ANI showed no significant changes in Dc, Bc, NCp, Ne, NLe, and NLp (all *P* > 0.05, Bonferroni correction).

### Connectivity-based analysis

Analysis of the differences in FCN between the two groups showed that, compared to HC, ANI patients exhibited significant increases and decreases in connectivity in two subnetworks (all *P* < 0.05, NBS correction). The network showing increased functional connectivity involved 30 nodes and 39 edges, mainly including the bilateral precentral gyrus, bilateral olfactory cortex, bilateral lingual gyrus, right straight muscle, right superior occipital gyrus, bilateral fusiform gyrus, bilateral postcentral gyrus, left superior parietal gyrus, bilateral precuneus, bilateral paracentral lobule, bilateral caudate nucleus, left putamen, right thalamus, left inferior temporal gyrus, right Cerebelum_Crus2, right Cerebelum_3, bilateral Cerebelum_4_5, left Cerebelum_7b, left Cerebelum_8, and Vermis_9. Overall, the subnetworks with increased functional connectivity in patients were mostly located in the control-executive network and visual network of functional partitions. The network showing decreased functional connectivity involved 19 nodes and 24 edges, mainly including the left superior frontal gyrus, left anterior cingulate and paracingulate gyrus, right parahippocampal gyrus, bilateral caudate nucleus, bilateral putamen, right pallidum, left Cerebelum_Crus1, bilateral Cerebelum_Crus2, right Cerebelum_3, left Cerebelum_4_5, left Cerebelum_7b, bilateral Cerebelum_8, left Cerebelum_9, Vermis_8, and Vermis_9. Overall, the subnetworks with decreased functional connectivity in ANI patients were mostly located in the default mode network (DMN) of functional partitions (left superior frontal gyrus, left anterior cingulate and paracingulate gyrus, and right parahippocampal gyrus), and the control-executive network (basal ganglia, cerebellum). Furthermore, structural connectivity analysis showed no significant differences between the two groups (*P* > 0.05, NBS correction). See Figure 4.

**Fig 4.**
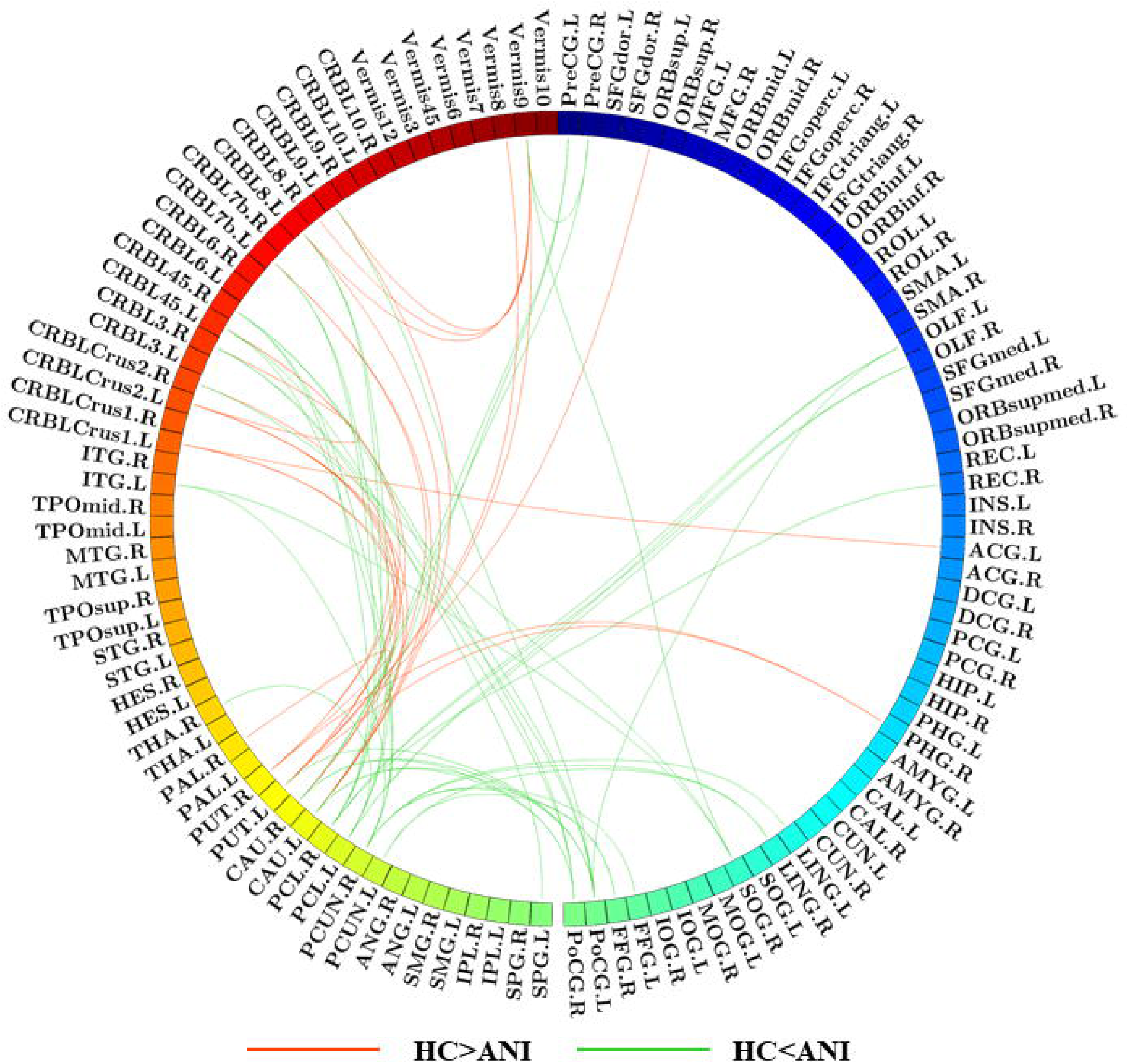
Functional network connectivity analysis between HC and ANI, with red indicating negative T-scores, suggesting reduced subnetwork connectivity in ANI, and green indicating positive T-scores, reflecting enhanced subnetwork connectivity in the ANI group (*P* < 0.05, NBS correction).

### Disrupted functional connectivity and structural connectivity network coupling

At the whole-brain connectivity level, no significant difference was found in the correlation coefficients between the HC and ANI patient groups (*P* = 0.083). At the modular level, ANI patients exhibited significantly enhanced functional-structural coupling in the prefrontal and occipital networks (P = 0.022 and P = 0.018), while no significant difference was observed between the two groups in the cerebellar network (P = 0.458). See Figure S2.

### Network Metrics in Relation to Clinical Variables and Cognitive Performance

In the correlation analysis between network topology changes and clinical variables, after FDR correction, the following significant results were found at the node level: In ANI patients, the DC of the left superior occipital gyrus was significantly negatively correlated with infection duration (r = -0.289, *P* = 0.049); The Ne of the left and right pericentral cortex, left and right lingual gyrus, and left superior occipital gyrus were significantly negatively correlated with both infection duration and treatment duration (r = -0.412, *P* = 0.004; r = -0.309, *P* = 0.035; r = -0.401, *P* = 0.005; r = -0.399, *P* = 0.005; r = -0.329, *P* = 0.024; r = -0.384, *P* = 0.008; r = -0.297, *P* = 0.043; r = -0.376, *P* = 0.009; r = -0.402, *P* = 0.005; r = -0.306, *P* = 0.036); In addition, the NLp of the left and right lingual gyrus was significantly positively correlated with both infection duration and treatment duration (r = 0.393, *P* = 0.006; r = 0.389, *P* = 0.007; r = 0.372, *P* = 0.01; r = 0.394, *P* = 0.006); The Ne of the left Cerebelum_6 was significantly positively correlated with both CD4^+^ cell count and its lowest value (r = 0.318, *P* = 0.029; r = 0.307, *P* = 0.036), while NLp was significantly negatively correlated with both CD4^+^ cell count and its lowest value (r = -0.321, *P* = 0.028; r = -0.293, *P* = 0.045); Furthermore, no significant correlation was found between network metrics and clinical variables in the analysis of small-world characteristics, global brain efficiency, and modular coupling. See Figure 5a.

**Fig 5.**
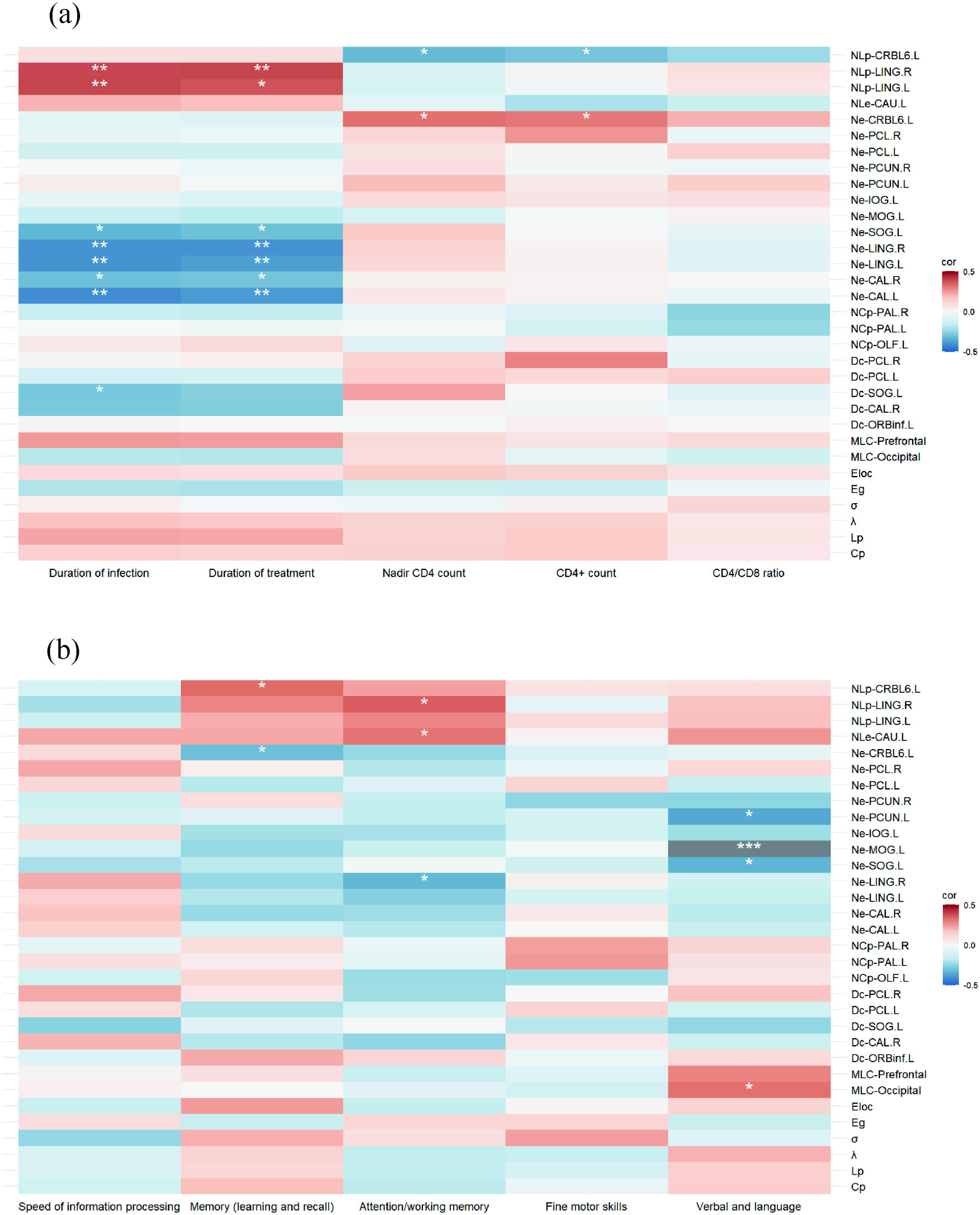
(a) Heatmap of the correlation analysis between changes in network topology and clinical variables. (b) Heatmap of the correlation analysis between changes in network topology and cognitive performance. * 0.01 < *P* ≤ 0.05, ** 0.001 < *P* ≤ 0.01, *** *P* ≤ 0.001. Abbreviations: MLC, modular-level coupling.

In the correlation analysis between network topology changes and cognitive performance, after FDR correction, the following significant results were found at the node level: The Ne of the left Cerebelum_6 was significantly negatively correlated with Memory (learning and recall) (r = -0.309, *P* = 0.035), while NLp was significantly positively correlated with it (r = 0.331, *P* = 0.023); The Ne of the right lingual gyrus was significantly negatively correlated with Attention/working memory (r = -0.323, *P* = 0.027), while NLp was significantly positively correlated with it (r = 0.352, *P* = 0.015); The NLe of the left caudate nucleus was significantly positively correlated with Attention/working memory (r = 0.312, *P* = 0.033); The Ne of the left superior occipital gyrus, left middle occipital gyrus, and left precuneus was significantly negatively correlated with Verbal and language (r = -0.334, *P* = 0.022; r = -0.507, *P* < 0.001; r = -0.361, *P* = 0.013). In the analysis of coupling at the modular level with cognitive performance, the functional-structural coupling of the occipital network was significantly positively correlated with Verbal and language (r = 0.32, *P* = 0.028). Moreover, no significant correlation was found between small-world characteristics, global brain efficiency, and cognitive performance. See Figure 5b.

## Discussion

This study focuses on the early stage of HAND, specifically the ANI phase, from the perspective of large-scale brain networks. We explored the characteristics of its functional and structural networks and the changes in functional-structural coupling. The findings indicate that although the structural network in ANI patients is relatively stable, significant reorganization of the functional network occurs in several regions (such as the visual network, executive control network, and default mode network), which is manifested by enhanced small-world properties and increased within-module connectivity. This suggests that, at the ANI stage, synaptic and dendritic damage at the microscopic level may first manifest in the functional network, serving as an "early warning signal" in the progression of HAND. Furthermore, functional-structural coupling is significantly enhanced in the occipital and frontal networks, suggesting that these regions play a core role in early neural damage. At the same time, changes in the topological characteristics and coupling of the functional network are closely associated with the patient’s immune status (such as CD4^+^ cell count) and cognitive performance. Therefore, the changes in functional network and coupling characteristics revealed by this study provide an objective basis for the early diagnosis and intervention of HAND, offering significant clinical application value.

### 1. Stability of Structural Networks in the Early Stage of HAND

Studies have shown that both the global and local properties of the structural network in ANI patients remain stable, indicating that the anatomical connections in the early stage of HAND are not yet significantly damaged. This is consistent with the pathological features of the ANI stage, such as reduced synaptic density and dendritic simplification^[52]^. Synaptic and dendritic damage in mild HAND (including ANI and MND) is primarily caused by inflammatory responses, oxidative stress, and direct HIV viral attack on neurons^[53, 54]^. This manifests as an imbalance in neurotransmitter release, including glutamate accumulation and insufficient dopamine secretion^[55–57]^. Glutamate excitotoxicity can lead to abnormal synaptic firing, thereby affecting neural activity^[10, 12]^, while dopamine system dysfunction alters the connectivity of the fronto-striatal circuit^[56, 58, 59]^. In this context, the reorganization of the functional network may be an early compensatory response to synaptic and dendritic damage^[60]^. Additionally, the brain may delay structural network damage by enhancing the efficiency of remaining connections. This compensatory mechanism has also been observed in other neurodegenerative diseases (such as Alzheimer’s disease and Parkinson’s disease)^[61, 62]^. However, this compensatory ability is limited. As the disease progresses, microscopic damage gradually accumulates, ultimately leading to the destruction of the macroscopic structural network. In the HAD stage, pathological features include a significant reduction in neurons in cortical and subcortical regions^[63–66]^, as well as gray matter atrophy and disruption of white matter integrity.

Therefore, the microscopic synaptic and dendritic damage in the ANI stage may first manifest in the functional network, serving as an "early warning signal" of the early stages of HAND. This finding not only provides clues for the early diagnosis of HAND but also opens new directions for exploring the dynamic changes in the functional-structural relationship.

### 2. Reorganization of Functional Networks

#### 2.1. Reorganization of the Visual Network (Occipital Lobe, Precuneus, Parahippocampal Cortex, Lingual Gyrus)

Small-world architecture and modular networks are key to the brain’s efficient information processing^[13, 67, 68]^. This study found that the functional networks of ANI patients exhibited a significant increase in small-world properties (σ↑, Eg↑, Lp↓), but lower local clustering (Cp↓) and local efficiency (Eloc↓). This suggests that while global network efficiency is enhanced, the local network’s information processing capacity may be limited. This could be related to early adaptive compensation triggered by synaptic and dendritic damage^[60]^. At the modular level of the visual network, ANI patients showed significantly enhanced intramodule connectivity in the occipital region. Additionally, node-level analysis revealed increased Dc in the visual network (occipital lobe, precuneus, parahippocampal cortex, lingual gyrus), but decreased Ne. This suggests that the information transmission efficiency in the occipital region may be impaired, and to compensate for this damage, the region may increase the number of network connections.

Previous studies have shown that the occipital lobe is one of the regions with the highest permeability to HIV Tat protein^[69]^. Its damage manifests in metabolic abnormalities^[52, 70]^, blood flow perfusion^[71]^, structural changes (posterior cortical gray matter atrophy^[72–74]^ and white matter integrity disruption^[75]^), and functional abnormalities (neural dynamics^[76, 77]^ and functional connectivity^[78–81]^). These damages often begin during the asymptomatic phase of infection and persist during cART treatment^[82]^. Therefore, the visual network in ANI patients enhances global efficiency by increasing intramodule connectivity, but the decline in local information processing capacity may reflect the limitations of the compensatory mechanism. This adaptive reorganization is highly consistent with the metabolic and structural abnormalities in the occipital region, further highlighting the key role of the occipital region in HAND.

#### 2.2. Functional Reorganization of the Control-Executive Network (Prefrontal Cortex, Cerebellum, Basal Ganglia)

This study found weakened intermodule (prefrontal and cerebellar network) connectivity in the control-executive network during the ANI stage. At the node level, significant reductions in Dc of the ORBinf.L and NLp in the CRBL6.L were observed, indicating impaired functional processing in these regions. The prefrontal cortex, a key region for higher-order cognitive processes, has been repeatedly identified as a site highly vulnerable to HIV-1 infection^[72, 83, 84]^. Post-mortem studies have shown that changes in synaptic structure in the prefrontal cortex of HIV-infected individuals are significantly associated with HAND^[85]^. Simultaneously, functional connectivity analysis revealed compensatory adaptation manifested as local activation or suppression in the cerebellar region. Additionally, Ne in the CRBL6.L was significantly positively correlated with CD4^+^ cell count and its lowest value, while NLp showed a significant negative correlation. Ne of the Cerebellum_6 was significantly negatively correlated with memory (learning and recall), while NLp showed a significant positive correlation. This suggests that the functional state of the Cerebellum_6 is not only closely related to immune status but also significantly influences cognitive performance. However, this region may face challenges in compensatory functional processing during information processing. This indicates that the cerebellum serves as a bridge between immune status and cognitive function, but its compensatory capacity is limited.

In the basal ganglia, the ANI group showed significant reductions in NCp in the bilateral globus pallidus and NLe in the left caudate nucleus. Functional connectivity analysis revealed activation or suppression within the basal ganglia (caudate nucleus, putamen, globus pallidus) in the ANI group, indicating basal ganglia dysfunction, leading to weakened local network connections and impaired information processing. The basal ganglia is a region highly vulnerable to HIV^[86–90]^, with elevated viral load and significant atrophy^[87, 91, 92]^. Specifically, the caudate nucleus, due to its proximity to the lateral ventricles, is vulnerable to HIV invasion through cerebrospinal fluid^[90, 93]^. Additionally, the high density of dopaminergic terminals in the striatum makes it particularly sensitive to HIV^[86]^, and dopamine plays a crucial role in higher cognitive functions such as memory, learning recognition, and attention shifting^[94]^. This study found that NLe in the left caudate nucleus of ANI patients was significantly positively correlated with attention and working memory (r = 0.312, *P* = 0.033). This suggests that HIV may affect these cognitive functions by reducing dopamine levels in the caudate nucleus.

Therefore, the multi-regional functional abnormalities in the executive network during the ANI stage exhibit features of adaptive reorganization, but the compensatory capacity is limited and cannot fully offset the weakened intermodule connectivity and decreased local information processing capacity.

#### 2.3. Reorganization of the DMN Network (Bilateral Fusiform Gyrus, Left Inferior Temporal Gyrus, Left Superior Orbital Gyrus, Left Anterior Cingulate and Paracingulate Gyrus)

FCN analysis revealed enhanced connectivity in the bilateral fusiform gyrus and left inferior temporal gyrus in ANI patients, while connectivity was reduced in the left superior orbital gyrus, left anterior cingulate, and paracingulate gyrus. These regions belong to the DMN in functional parcellation, and DMN is closely associated with higher-order cognitive functions such as self-reflection, emotional regulation, and memory retrieval^[95]^. Previous studies have suggested that reorganization of the DMN in HAND may be an adaptation to local damage, but it could also represent potential impairment of network integration capabilities^[81, 96, 97]^.

### 3. Abnormalities in Structure-Function Coupling

In ANI patients, the function-structure coupling in the occipital and frontal networks was significantly enhanced, suggesting that these brain regions may maintain functional stability by strengthening the synergy between function-structure connections in response to early neural damage. Specifically, the function-structure coupling of the occipital network was significantly enhanced in ANI patients, further supporting the adaptive response of this network to neural damage. Moreover, the node efficiency of the left superior occipital gyrus, middle occipital gyrus, and precuneus was negatively correlated with the verbal and language domain, whereas the function-structure coupling of the occipital network was positively correlated with verbal and language performance. This suggests that the decline in visual and spatial processing abilities may affect language performance, while also reflecting the network’s adaptive capacity in complex cognitive tasks. In the right lingual gyrus, the Ne was significantly negatively correlated with attention/working memory, while the NLp was positively correlated. This suggests that the lingual gyrus may compensate by increasing path connectivity, thereby maintaining functional integrity in complex cognitive tasks. Additionally, the Ne and Np of the bilateral cuneal cortical regions and lingual gyrus in the occipital network were correlated with infection and treatment duration. This further suggests that the adaptability of the occipital network in HIV infection is limited, and may gradually decline as the infection persists.

The significant enhancement of the function-structure coupling in the prefrontal network in the ANI group may represent a compensatory mechanism for the weakened connectivity between the prefrontal and cerebellar network modules. Previous studies have indicated that there may be strong functional connectivity between the prefrontal cortex and the cerebellum_6^[98]^, and functional connectivity abnormalities between the frontal, parietal, and cerebellar networks have been repeatedly reported in HAND patients^[99, 100]^. Although no significant changes in the cerebellar network’s function-structure coupling were observed in this study, the functional connectivity network indicates that cerebellar regions may still retain some functional activity in the ANI phase. Despite being damaged, the cerebellar network still exhibits some degree of adaptability, striving to maintain its functional integrity through function-structure coupling, but this compensatory capacity may be limited.

Our study has several limitations. Firstly, the small sample size limits the generalizability of the observed changes in the ANI group. Therefore, these findings need to be validated in larger cohorts to confirm their applicability. Secondly, all participants were male, which may introduce a gender bias. Recent studies suggest that there may be gender differences in the incidence and patterns of neurocognitive disorders^[101]^.

This study reveals key neural network characteristics of the early stage of HAND, specifically during the ANI phase, from a multimodal brain network perspective. It finds that while the structural network remains stable, the functional network undergoes significant reorganization, characterized by enhanced small-world properties and increased within-module connectivity.

These changes may represent early compensatory signals for synaptic and dendritic damage. The significant enhancement of function-structure coupling in the occipital and frontal networks reflects the critical role of these regions in maintaining brain functional stability. The topological characteristics and coupling changes in the functional network are closely related to the patients’ immune status and cognitive performance. This study provides important scientific evidence for the precise diagnosis, personalized treatment, and management of HAND.

## Acknowledgments

This study expresses gratitude to those who previously contributed to the project, particularly in data collection, even though some researchers are no longer active in the field of HIV-associated neurocognitive disorders. Their contributions remain invaluable. These individuals include Dan Liu, Yu Qi, Yuxun Gao, Shuai Han, and Yuanyuan Wang, to whom we extend our sincere thanks.

## Author contribution

Author contributions included conception and study design (ZZ and WW), data collection or acquisition (ZZ, WG and FW), statistical analysis (ZZ, HH and HL), interpretation of results (ZZ, WW, WG and FX), drafting the manuscript work or revising it critically for important intellectual content (ZZ, WW, HL, WG, HH, FW, HL and FX) and approval of final version to be published and agreement to be accountable for the integrity and accuracy of all aspects of the work (All authors).

## Data availability

The data that support the findings of this study are available upon reasonable request.

## Declarations

### Ethics approval

This study was approved by the Medical Ethics Committee of Beijing Youan Hospital, Capital Medical University (Ethics Approval No. LL-2020-047-K).

### Consent to participate

Written informed consent was obtained from all study participants.

### Consent to publish

All authors consented this paper to be published in Brain Imaging and Behavior.

