## Supplementary material for "Large-scale brain network analysis reveals functional-structural dissynchrony in HIV-associated asymptomatic neurocognitive disorders: Functional disturbances precede structural changes": Table S1, Table S2, Table S3,Table S4, Figure S1, Table S5, Figure S2

**Appendices**

**Content**

Supplementary table 1: Regions of interest (ROI) in the AAL template………………2

Supplementary table 2: Demographic, clinical and cognitive characteristics of the participants………………………………….……………………………….………...5

Supplementary table 3: Alterations in global functional network metrics are observed in ANI patients……………………………………………………………………..….6

Supplementary table 4: Altered node attributes were observed in the functional brain networks of ANI patients……………………………………………………………....7

Supplementary figure 1: Structural network global properties remain stable in ANI patients………………………………………………………………………………...9

Supplementary table 5: Structural network global properties remain stable in ANI patients……………………………………………………………………………….10

Supplementary figure 2: Disrupted functional connectivity and structural connectivity network coupling……………………………………………………………………..11

**Table S1**

Regions of interest (ROI) in the AAL template

| Region name | Abbreviation |
| --- | --- |
| Precentral | PreCG |
| Superior frontal gyrus (dorsolateral) | SFGdor |
| Superior frontal gyrus (orbital part) | ORBsup |
| Middle frontal gyrus | MFG |
| Middle frontal gyrus (orbital part) | ORBmid |
| Inferior frontal gyrus (opercular part) | IFGoperc |
| Inferior frontal gyrus (triangular part) | IFGtriang |
| Inferior frontal gyrus (orbital part) | ORBinf |
| Rolandic operculum | ROL |
| Supplementary motor area | SMA |
| Olfactroy cortex | OLF |
| Superior frontal gyrus (medial) | SFGmed |
| Superior frontal gyrus (medial orbital) | ORBsupmed |
| Rectus gyrus | REC |
| Insula | INS |
| Anterior cingulate gyri | ACG |
| Median cingulate gyri | MCG |
| Posterior cingulate gyrus | PCG |
| Hippocampus | HIP |
| Parahippocampal gyrus | PHG |
| Amygdala | AMYG |
| Calcarine fissure | CAL |
| Cuneus | CUN |
| Lingual gyrus | LING |
| Superior occipital gyrus | SOG |
| Middle occipital gyrus | MOG |
| Inferior occipital gyrus | IOG |
| Fusiform gyrus | FFG |
| Postcentral gyrus | PoCG |
| Superior parietal gyrus | SPG |
| Inferior parietal gyrus | IPG |
| Supramarginal gyrus | SMG |
| Angular gyrus | ANG |
| Precuneus | PCUN |
| Paracentral lobule | PCL |
| Caudate nucleus | CAU |
| Putamen | PUT |
| Pallidum | PAL |
| Thalamus | THA |
| Heschl gyrus | HES |
| Superior temporal gyrus | STG |
| Superior temporal gyrus, temporal pole | TPOsup |
| Middle temporal gyrus | MTG |
| Middle temporal gyrus, temporal pole | TPOmid |
| Inferior temporal gyrus | ITG |
| Cerebelum_Crus1 | N/A |
| Cerebelum_Crus2 | N/A |
| Cerebelum_3 | N/A |
| Cerebelum_4_5 | N/A |
| Cerebelum_6 | N/A |
| Cerebelum_7b | N/A |
| Cerebelum_8 | N/A |
| Cerebelum_9 | N/A |
| Cerebelum_10 | N/A |
| Vermis_1_2 | N/A |
| Vermis_3 | N/A |
| Vermis_4_5 | N/A |
| Vermis_6 | N/A |
| Vermis_7 | N/A |
| Vermis_8 | N/A |
| Vermis_9 | N/A |
| Vermis_10 | N/A |

The left and right brain labels are combined in the display, whereas cerebellar acronyms are not applicable.

**Table S2**

Demographic, clinical and cognitive characteristics of the participants

| Variable | HC (n = 48) | ANI (n = 47) | *P*-value |
| --- | --- | --- | --- |
| Age (years) | 0.742 ± 0.020 | 0.741 ± 0.022 | 0.844 ^a^ |
| Male, n (%) | 47 (97.9%) | 47 (100.0%) | 0.320 ^b^ |
| Education years (years) | 14.792 ±3.274 | 14.638 ±2.937 | 0.811 ^a^ |
| Duration of infection (months) | N/A | 53.064 ± 29.723 | N/A |
| Duration of treatment (months) | N/A | 49.957 ± 29.429 | N/A |
| Nadir CD4^+^ count (cells/μL) | N/A | 373.064 ± 201.815 | N/A |
| CD4^+^ count (cells/μL) | N/A | 565.426 ± 242.456 | N/A |
| CD4/CD8 radio | N/A | 0.680 ± 0.325 | N/A |
| TND, n (%) | N/A | 47 (100.0%) | N/A |
| Speed of information processing | 50.771 ± 7.096 | 41.532 ± 9.899 | < 0.001 ^a^ |
| Memory (learning and recall) | 49.797 ± 5.482 | 31.575 ± 7.750 | < 0.001 ^a^ |
| Attention/working memory | 51.844 ± 6.306 | 32.053 ± 9.018 | < 0.001 ^a^ |
| Fine motor skills | 51.750 ± 5.930 | 41.149 ± 9.818 | < 0.001 ^a^ |
| Verbal and language | 56.292 ± 6.737 | 46.596 ± 10.147 | < 0.001 ^a^ |

Statistical tests including: ^a^*t*-test; ^b^chi-square test. Abbreviations: TND, virus not detectable.

**Table S3**

Alterations in global functional network metrics are observed in ANI patients

| Global network measures | HC group (n = 48) | PT group (n = 47) | *t*-value | *P*-value |
| --- | --- | --- | --- | --- |
| Cp | 0.260 ± 0.033 | 0.245 ± 0.013 | 2.921 | 0.004 |
| γ | 1.011 ± 0.071 | 1.006 ± 0.067 | 0.340 | 0.735 |
| λ | 0.498 ± 0.050 | 0.479 ± 0.007 | 2.687 | 0.009 |
| Lp | 0.848 ± 0.110 | 0.803 ± 0.018 | 2.761 | 0.007 |
| σ | 0.889 ± 0.072 | 0.918 ± 0.058 | -2.180 | 0.032 |
| Eg | 0.260 ± 0.017 | 0.267 ± 0.003 | -2.843 | 0.005 |
| Eloc | 0.344 ± 0.017 | 0.337 ± 0.007 | 2.903 | 0.005 |

Positive and negative t-values indicate the direction of the effect in the statistical test.

**Table S4**

Altered node attributes were observed in the functional brain networks of ANI patients

| Brain regions | HC group (n = 48) | PT group (n = 47) | *t*-value | *P*-value |
| --- | --- | --- | --- | --- |
| Dc  ORBinf.L | 15.589 ± 3.617 | 13.189 ± 2.348 | 3.828 | 0.00023 |
| CAL.R | 11.244 ± 2.618 | 13.406 ± 2.458 | -4.148 | 0.00007 |
| SOG.L | 11.500 ± 2.787 | 13.392 ± 1.852 | -3.888 | 0.00019 |
| PCL.L | 12.579 ± 2.622 | 14.796 ± 2.445 | -4.261 | 0.00005 |
| PCL.R | 11.813 ± 2.386 | 14.050 ± 2.534 | -4.430 | 0.00003 |
| NCp |  |  |  |  |
| OLF.L | 0.258 ± 0.036 | 0.231 ± 0.035 | 3.689 | 0.00038 |
| PAL.L | 0.262 ± 0.056 | 0.235 ± 0.038 | 3.666 | 0.00041 |
| PAL.R | 0.256 ± 0.049 | 0.234 ± 0.041 | 3.672 | 0.00040 |
| Ne |  |  |  |  |
| CAL.L | 0.246 ± 0.021 | 0.262 ± 0.016 | 4.507 | 0.00002 |
| CAL.R | 0.242 ± 0.026 | 0.262 ± 0.015 | 4.465 | 0.00002 |
| LING.L | 0.254 ± 0.016 | 0.266 ± 0.015 | 3.695 | 0.00037 |
| LING.R | 0.253 ± 0.016 | 0.268 ± 0.015 | 4.578 | 0.00001 |
| SOG.L | 0.242 ± 0.033 | 0.265 ± 0.013 | 3.933 | 0.00016 |
| MOG.L | 0.247 ± 0.030 | 0.265 ± 0.013 | 3.759 | 0.00030 |
| IOG.L | 0.251 ± 0.017 | 0.263 ± 0.011 | 3.981 | 0.00014 |
| PCUN.L | 0.245 ± 0.033 | 0.265 ± 0.013 | 3.916 | 0.00017 |
| PCUN.R | 0.246 ± 0.033 | 0.268 ± 0.014 | 3.918 | 0.00017 |
| PCL.L | 0.247 ± 0.035 | 0.270 ± 0.016 | 4.131 | 0.00008 |
| PCL.R | 0.242 ± 0.034 | 0.267 ± 0.014 | 4.457 | 0.00002 |
| CRBL6.L | 0.270 ± 0.016 | 0.282 ± 0.014 | 3.864 | 0.00021 |
| NLe |  |  |  |  |
| CAU.L | 0.270 ± 0.023 | 0.264 ± 0.016 | 3.842 | 0.00022 |
| NLp |  |  |  |  |
| LING.L | 0.848 ± 0.073 | 0.793 ± 0.049 | 4.364 | 0.00003 |
| LING.R | 0.855 ± 0.074 | 0.789 ± 0.047 | 5.165 | 0.00001 |
| CRBL6.L | 0.800 ± 0.071 | 0.748 ± 0.043 | 4.386 | 0.00003 |

Positive and negative t-values indicate the direction of the effect in the statistical test.

**Figure S1**

Structural network global properties remain stable in ANI patients

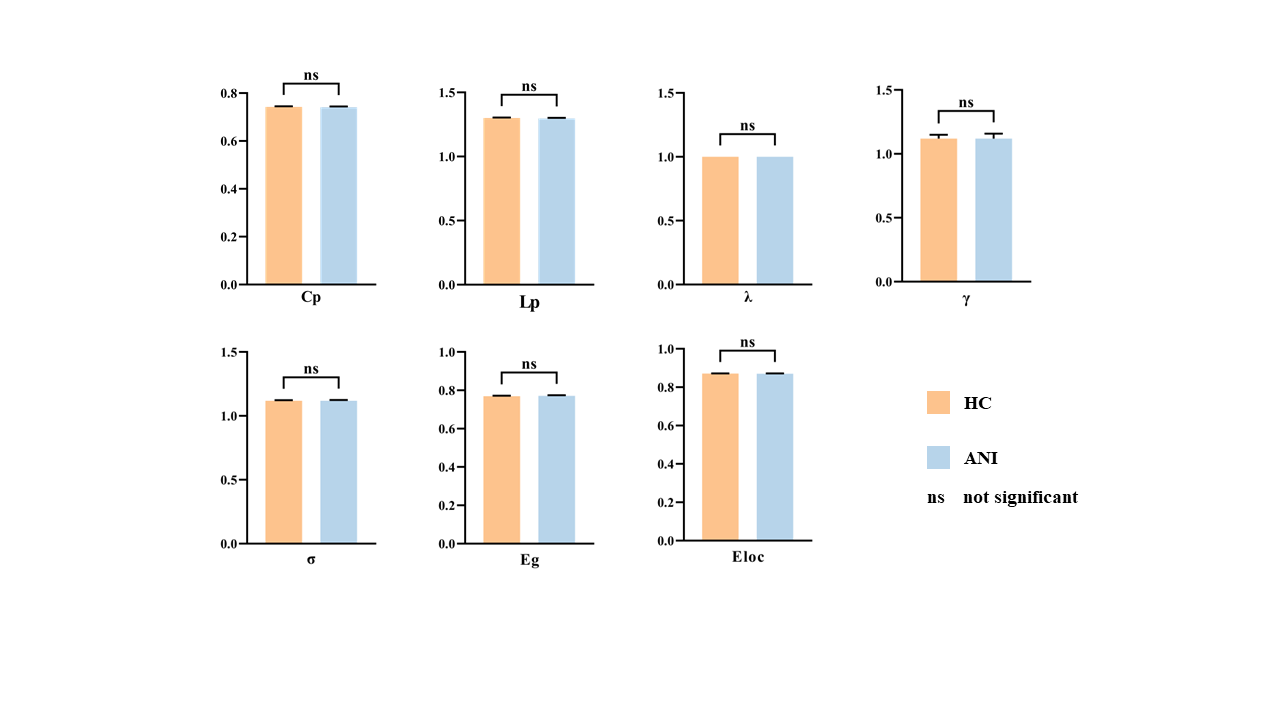
There were no significant differences in the global properties of the structural connectivity network between HC and ANI. The bar chart represents the mean values of each metric for the subjects, with vertical bars indicating the standard error between subjects. Abbreviations: ns, not significant.

**Table S5**

Structural network global properties remain stable in ANI patients

| Global network measures | HC group (n=48) | PT group (n = 47) | *t*-value | *P*-value |
| --- | --- | --- | --- | --- |
| Cp | 0.742 ± 0.020 | 0.741 ± 0.022 | 0.197 | 0.844 |
| γ | 1.120 ± 0.031 | 1.120 ± 0.039 | -0.004 | 0.997 |
| λ | 1.001 ± 0.001 | 1.001 ± 0.001 | 1.510 | 0.134 |
| Lp | 1.300 ± 0.031 | 1.300 ± 0.035 | 0.485 | 0.629 |
| σ | 1.118 ± 0.030 | 1.118 ± 0.038 | -0.031 | 0.976 |
| Eg | 0.770 ± 0.018 | 0.772 ± 0.021 | -0.511 | 0.610 |
| Eloc | 0.871 ± 0.010 | 0.871 ± 0.011 | 0.197 | 0.845 |

Positive and negative t-values indicate the direction of the effect in the statistical test.

**Figure S2**

Disrupted functional connectivity and structural connectivity network coupling

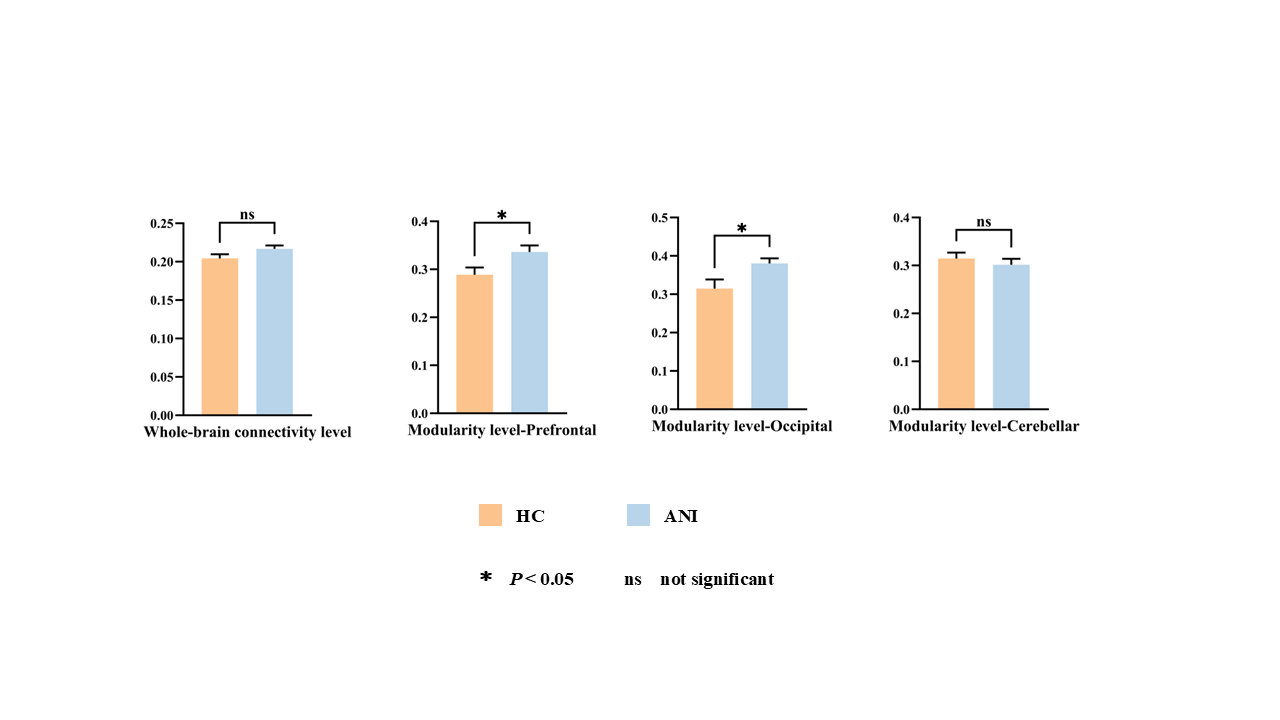

Compared to HC, there were no changes in the functional-structural connectivity coupling at the whole-brain level in ANI patients. However, at the modular level, the functional-structural coupling between the prefrontal and occipital networks was enhanced in ANI patients.
